## Supplementary Appendix for "Beyond Borders: Spatial Disparities in the Mortality Burden of the Covid-19 pandemic across 569 European Regions (2020-2021)"

### **Online supplementary appendix A**

**Table A1: Regional division, sources, data information and adjustments by country**

| **Country** | **Spatial units** | **Source** | **Period** | **Upper age limit** | **Adjustments/comments** |
| --- | --- | --- | --- | --- | --- |
| Austria | 9 NUTS 2 units (‘Länder’) | Statistics Austria, Eurostat | 1990–2021 | Deaths: 115  Pop.: 95 (after 2001:100) | – |
| Belgium | 11 NUTS 2 units (‘provincies’) | Belgian Statistical Office | 1993–2021 | Deaths: 100  Pop.: 100 | – |
| Czechia | 14 NUTS 3 units (‘kraje’) | Czech Statistical Office | 1996–2021 | Deaths: 95  Pop.: 95 | – |
| Denmark | 11 NUTS 3 units (‘landsdele’) | Statistics Denmark | 2008–2020 | Deaths: 90  Pop.: 90 | – |
| Estonia | 1 NUTS 2 unit | Statistics Estonia | 1989– 2021 | Deaths:100  Pop.: 85 | – |
| Finland | 4 NUTS 2 units  (‘Storområden’) | Statistics Finland | 1972– 2021 | Deaths: 95  Pop.: 95 | We merged Åland Islands to South Finland |
| France | 95 NUTS 3 units (‘départements’) | INSEE | 1970–2021 | Deaths: 105  Pop.: 105 (after 2016: 95) | Non-European areas excluded; north and south of Corse merged to maintain consistent time series |
| Germany | 96 ROR (‘Raumordnungs-regionen’) | Statistical Offices of the German Länder | 1992–2021 | Deaths: 90  Pop.: 90 | Harmonized to apply current territorial administrative division (as of Dec 2022) to the whole study period and to eliminate the Census 2011 break; 400 NUTS 3 units (‘Kreise’) aggregated to 96 ROR units according to classification of BBSR (2017) [36] |
| Hungary | 8 NUTS 2 units (‘tervezési-statisztikai régiók’) | Hungarian Central Statistical Office, Eurostat | 2001–2021 | Deaths: 90  Pop.: 90 | – |
| Iceland | 1 NUTS 2 unit | Human Mortality Database, Eurostat | 1970–2021 | Deaths: 110 (2021:100)  Pop.: 110 (2021:100) | – |
| Ireland | 1 NUTS 1 unit | Human Mortality Database, Eurostat | 1990–2021 | Deaths: 110 (2021:100)  Pop.: 110 (2021:100) | No subnational division due to data availability issues |
| Italy | 92 NUTS 3 units (‘province’) | ISTAT | 1995–2021 | Deaths: 100  Pop.: 100 | We merged the following regions to maintain a consistent time series: 1) Biella + Vercelli, 2) Novara + Verbano, 3) Como + Lecco, 4) Milano + Lodi + Monza + Brianza, 5) Rimini + Forli-Cesena, 5) Firenze + Prato, 6) Cagliari + Medio Campidano + Carbonia-Iglesias + Ogliastra + Oristano + Nuoro, 7) Sassari + Olbia-Tempio, 8) Foggia + Bari + Barletta, 9) Fermo + Ascoli-Piceno, 10) Crotone + Vibo Valentia + Cantanzaro. |
| Latvia | 1 NUTS 2 unit | Official Statistics Portal Latvia | 1990–2021 | Deaths: 100  Pop.: 100 | – |
| Lithuania | 2 NUTS 2 units | Statistical Office, Eurostat | 2001–2021 | Deaths: 85  Pop.: 100 | – |
| Luxembourg | 1 NUTS 3 unit | Human Mortality Database | 1996–2021 | Deaths: 110  Pop.: 110 | – |
| Netherlands | 12 NUTS 2 units (‘provincies’) | Statistics Netherlands | 1990–2021 | Deaths: 100  Pop.: 90 (after 2001: 100) | – |
| Norway | 7 NUTS 2 units (‘landsdeler’) | Statistics Norway | 2000–2021 | Deaths: 100  Pop.: 105 | Harmonized to apply current territorial administrative divisions; we excluded remote islands Svalbard and Jan Mayen |
| Poland | 73 NUTS 3 units (‘podregiony’) | Statistics Poland | 2006–2021 | Deaths: 90  Pop.: 100 | – |
| Portugal | 5 NUTS 2 units (‘regiões’) | National Institute of Statistics, Eurostat | 1992–2021 | Deaths: 100  Pop.: 100 | For visibility reasons, we excluded the remote islands Azores and Madeira. |
| Slovakia | 8 NUTS 3 units (‘kraje’) | Slovakian Statistical Office | 1996–2022 | Deaths: 100  Pop.: 100 | – |
| Slovenia | 2 NUTS 2 units (‘kohezijske regije’) | Slovenian Statistical Office | 2002–2021 | Deaths: 100  Pop.: 100 | – |
| Spain | 50 NUTS 3 units (‘provincias’) | National Statistics Institute | 1990–2021 | Deaths: 100  Pop.: 85 (after 2001:100) | For visibility reasons, we excluded the Canary Islands. |
| Sweden | 21 NUTS 3 units (‘län’) | Statistics Sweden, Eurostat | 1969–2021 | Deaths: 100  Pop.: 100 | – |
| Switzerland | 7 NUTS 2 units (‘Grossregionen’) | Federal Statistical Office | 1991–2020 | Deaths: 95  Pop.: 100 | – |
| United Kingdom | 37 NUTS 2 regions | Office for National Statistics (for England & Wales); Human Mortality Database (for Northern Ireland & Scotland) | 2002–2021  2002–2021  1990–2021 | England & Wales  Deaths: 90  Pop.: 90  Northern Ireland  Deaths: 110  Pop.: 110  Scotland  Deaths: 113  Pop: 90 | _  NUTS 1 level data for Northern Ireland and Scotland due to data quality issues |

**Table A2: Comparison of changes in life expectancy at birth with Schöley et al. (2022)**

|  | **Women** | | | | **Men** | | | |
| --- | --- | --- | --- | --- | --- | --- | --- | --- |
|  | **Schöley et al. 2022** | | **This paper** | | **Schöley et al. 2022** | | **This paper** | |
| **Country** | **2020** | **2021** | **2020** | **2021** | **2020** | **2021** | **2020** | **2021** |
| Austria | -0,70 | -0,70 | -0,49 | -0,59 | -0,99 | -1,26 | -0,81 | -1,12 |
| Belgium | -1,15 | -0,33 | -0,91 | -0,22 | -1,29 | -0,78 | -1,15 | -0,70 |
| Switzerland | -0,64 | -0,23 | -0,90 | -0,55 | -1,14 | -0,74 | -1,57 | -1,17 |
| Czech Republic | -0,94 | -1,76 | -0,94 | -1,94 | -1,25 | -2,41 | -1,20 | -2,60 |
| Germany | -0,32 | -0,67 | -0,10 | -0,52 | -0,49 | -1,03 | -0,30 | -0,84 |
| Denmark | -0,03 | -0,33 | 0,00 | -0,32 | -0,04 | -0,17 | -0,03 | -0,20 |
| Estonia | -0,32 | -1,94 | -0,07 | -1,51 | -0,40 | -2,65 | -0,23 | -2,12 |
| Spain | -1,41 | -0,93 | -1,14 | 0,02 | -1,52 | -1,22 | -1,38 | -0,44 |
| Finland | -0,01 | -0,23 | -0,02 | -0,24 | -0,30 | -0,32 | -0,37 | -0,47 |
| France | -0,52 | -0,18 | -0,43 | -0,46 | -0,75 | -0,53 | -0,71 | -0,85 |
| North. Ireland | -0,84 | -0,95 | -0,81 | -0,83 | -0,79 | -0,97 | -0,85 | -1,05 |
| Scotland | -0,45 | -0,69 | -0,44 | -0,73 | -1,07 | -1,00 | -0,91 | -0,83 |
| Hungary | -0,83 | -2,20 | -0,73 | -2,15 | -0,93 | -2,55 | -0,84 | -2,68 |
| Iceland | -0,41 | -0,39 | 0,01 | 0,01 | -0,30 | -0,11 | -0,22 | 0,04 |
| Italy | -1,07 | -0,97 | -0,86 | -0,76 | -1,46 | -1,25 | -1,33 | -1,12 |
| Lithuania | -1,57 | -2,97 | -1,08 | -2,43 | -2,25 | -2,98 | -1,69 | -2,58 |
| Netherlands | -0,58 | -0,74 | -0,52 | -0,76 | -0,95 | -0,94 | -0,90 | -1,10 |
| Norway | 0,04 | -0,21 | -0,05 | -0,19 | -0,05 | -0,17 | -0,26 | -0,26 |
| Poland | -1,02 | -2,26 | -1,02 | -2,08 | -1,60 | -2,64 | -1,55 | -2,55 |
| Portugal | -0,66 | -0,65 | -0,59 | -0,64 | -0,91 | -1,02 | -0,77 | -0,78 |
| Sweden | -0,62 | -0,21 | -0,30 | 0,02 | -1,01 | -0,56 | -0,72 | -0,31 |
| Slovenia | -1,02 | -0,64 | -0,87 | -0,77 | -0,93 | -0,88 | -1,00 | -1,38 |
| Slovakia | -0,94 | -2,97 | -0,85 | -3,09 | -1,10 | -3,38 | -1,07 | -3,58 |

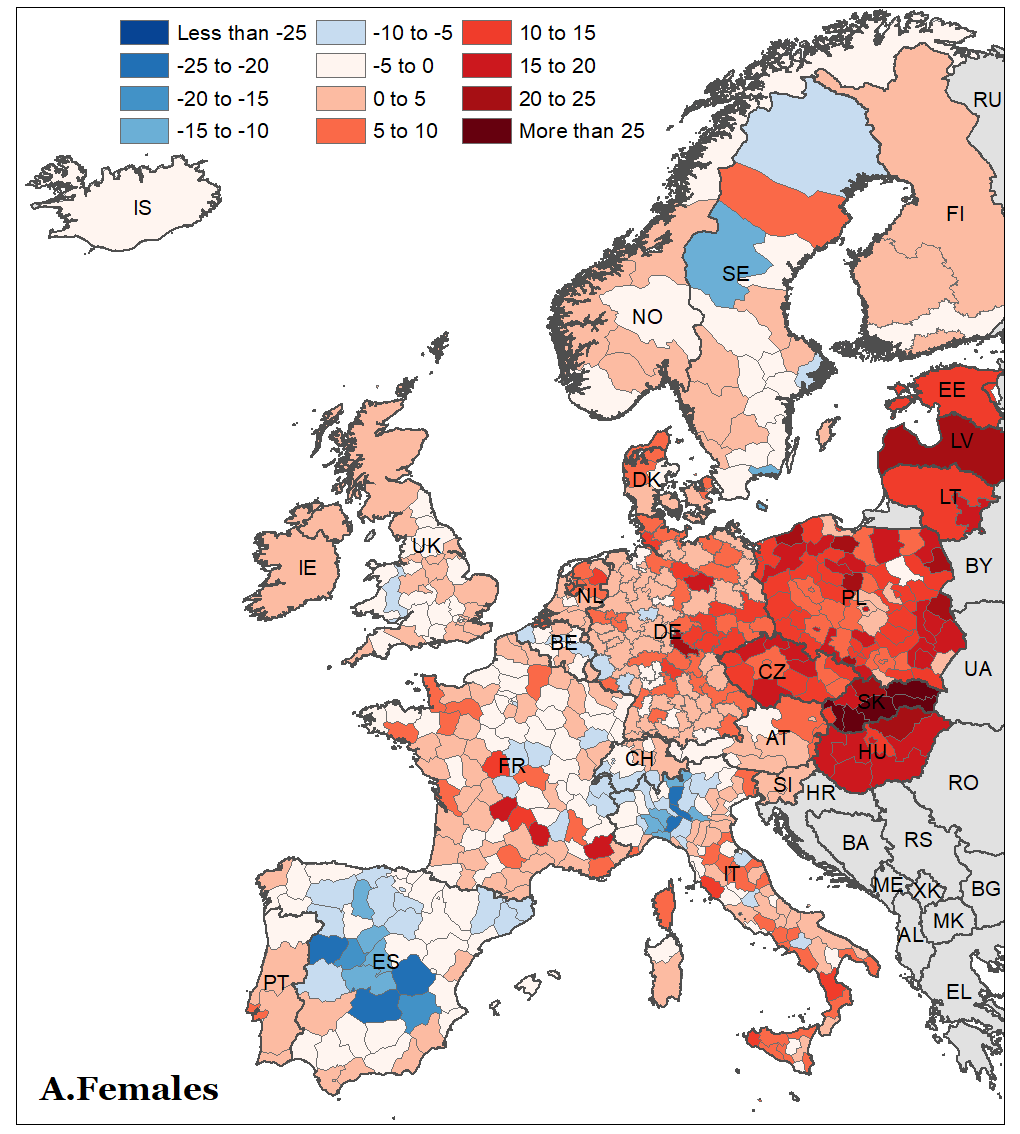

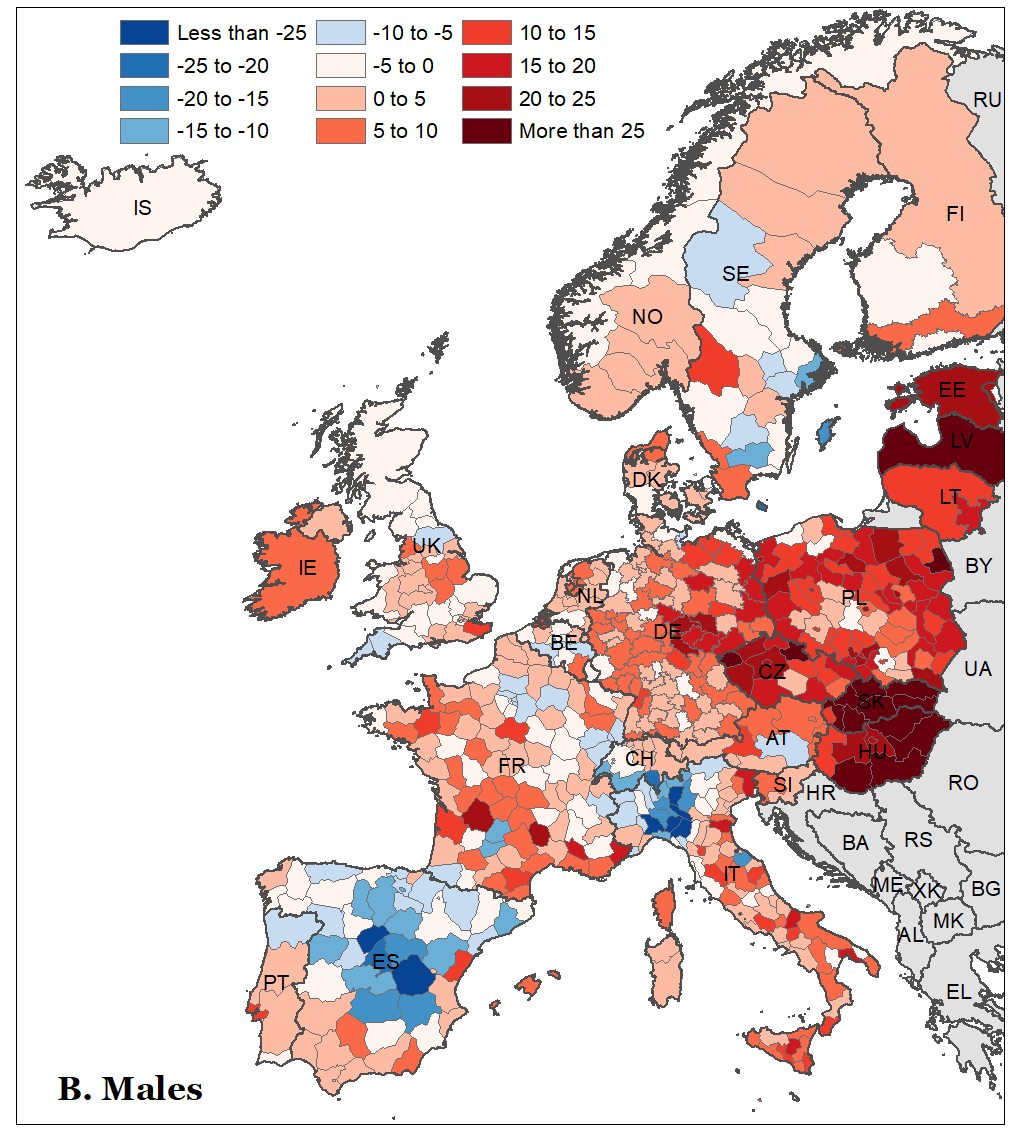

**Figure A1. Spatial distribution of the change in age-standardized years of life lost (ASYLL) between 2020 and 2021 across 25 European countries**

Source: As shown in Table A1

### **Online supplementary appendix B**

A detailed description of the analytic procedure to compute excess mortality is available at:

<https://osf.io/fwtsa/?view_only=ba00308358dc4fbaa23de72f9c82d1db>

### **Online supplementary appendix C**

Detailed values of our estimates and data visualisation tool are available at:

<https://osf.io/fwtsa/?view_only=ba00308358dc4fbaa23de72f9c82d1db>

Please read “Online Appendix C.pdf” first.
